# Understanding transmission risk and predicting environmental suitability for Mayaro Virus in the Americas

**DOI:** 10.1101/2023.04.13.23288376

**Authors:** Michael Celone, Sean Beeman, Barbara A. Han, Alexander M. Potter, David B. Pecor, Bernard Okech, Simon Pollett

## Abstract

**Background:** Mayaro virus (MAYV) is a mosquito-borne *Alphavirus* that is widespread in South America. MAYV infection often presents with non-specific febrile symptoms but may progress to debilitating chronic arthritis or arthralgia. Despite the pandemic threat of MAYV, its true distribution remains unknown. The objective of this study was to clarify the geographic distribution of MAYV using an established risk mapping framework. This consisted of generating evidence consensus scores for MAYV presence, modeling the potential distribution of MAYV across the Americas, and estimating at-risk population residing in areas suitable for MAYV transmission.

**Methods:** We compiled a georeferenced compendium of MAYV occurrence in humans, animals, and arthropods. Based on an established evidence consensus framework, we integrated multiple information sources to assess the total evidence supporting ongoing transmission of MAYV within each country in our study region. We then developed high resolution maps of the disease’s estimated distribution using a boosted regression tree approach. Models were developed using ten climatic and environmental covariates that are related to the MAYV transmission cycle. Using the output of our boosted regression tree models, we estimated the total population living in regions suitable for MAYV transmission.

**Findings:** The evidence consensus scores revealed high or very high evidence of MAYV transmission in Brazil (especially the states of Mato Grosso and Goiás), Venezuela, Peru, Trinidad and Tobago, Colombia, Bolivia, and French Guiana. According to the boosted regression tree models, a substantial region of South America is suitable for MAYV transmission, including north and central Brazil, French Guiana, and Suriname. Some regions (e.g., Guyana) with low or moderate evidence of transmission were identified as highly suitable for MAYV. We estimate that approximately 77 million people in the Americas live in areas that may be suitable for MAYV transmission, including 43·4 million people in Brazil. Our results can assist public health authorities in prioritizing high-risk areas for vector control, human disease surveillance and ecological studies.

**Funding:** This work was financially supported by the Armed Forces Health Surveillance Division—Global Emerging Infections Surveillance (AFHSD-GEIS) under awards P0065_22_WR and P0050_23_WR. The activities undertaken at WRBU were performed in part under a Memorandum of Understanding between the Walter Reed Army Institute of Research (WRAIR) and the Smithsonian Institution, with institutional support provided by both organizations.

## Research in context

### Evidence before this study

We searched PubMed on January 27, 2021 using the search term “Mayaro virus”. This search yielded 274 results including two systematic reviews of Mayaro virus (MAYV) occurrence and one study that modeled the distribution of MAYV risk. Although prior systematic reviews included some spatial information, they did not attempt to georeference MAYV occurrence with a high level of spatial precision and did not quantify uncertainty in geolocated records. Furthermore, the one published study that modeled environmental suitability for MAYV included only a limited set of MAYV occurrence locations. No additional studies have been conducted to model the risk of MAYV occurrence across space or to quantify the total population which may be at risk of MAYV infection in the Americas.

### Added value of this study

We used a comprehensive, georeferenced compendium of MAYV occurrence to model the suitability for MAYV occurrence in the Americas. Our boosted regression tree model incorporated 203 MAYV occurrence locations and 10 gridded environmental covariates to generate a 5 x 5 km continuous surface of MAYV suitability across the Americas. Using this distribution model, we estimated the total population residing in areas that are suitable for MAYV transmission. Furthermore, we developed evidence consensus scores for each country in our study region that synthesized a variety of sources to assess the overall evidence of MAYV transmission.

### Implications of all the available evidence

Our study provides a contemporary estimate of MAYV distribution using a well-established disease mapping framework. This information provides an evidence base that can guide disease surveillance (including human cases and ecological studies) and vector control efforts in the Americas. This is especially useful in regions with high MAYV suitability but little or no evidence of MAYV transmission (e.g., Guyana and Suriname).

## Introduction

Mayaro virus (MAYV) is a mosquito-borne *Alphavirus* that was first detected in Trinidad in 1954.^1^ MAYV has caused periodic outbreaks throughout Latin America^2^ and serological surveys and syndromic surveillance studies suggest widespread circulation in the region.^3^ Some researchers have hypothesized that MAYV has broader epidemic potential and raised alarm about its increased geographic spread.^4, 5^

MAYV can cause debilitating arthralgia or arthritis that can persist for months after initial infection.^6^ However, most MAYV patients present with non-specific febrile symptoms that are clinically indistinguishable from other vector borne diseases such as dengue or Zika.^7^ Therefore, clinical diagnosis is often difficult and accurate estimates of disease burden remain elusive. This is further complicated by the many limitations of serological diagnostics including the cross-reactivity of antigenically similar viruses.^8^ Supportive care remains the current standard of clinical treatment for MAYV as no licensed vaccine or antiviral treatment currently exists.

Limited studies on MAYV ecology suggest that this virus is maintained in a sylvatic transmission cycle involving arboreal mosquito vectors and non-human animal reservoirs. High seroprevalence among non-human primates (NHPs)^9^ suggests they may be involved in the MAYV transmission cycle, although their precise role is inconclusive. In addition, MAYV antibodies have been detected in other mammals including rodents and marsupials^10^. Risk factors including residing near forested areas^11^ and hunting in the rainforest^12^ are associated with MAYV infection in humans, highlighting the importance of the sylvatic transmission cycle and the potential for spillover events. However, the occurrence of MAYV in the city of Manaus has also led to concerns about the involvement of *Aedes (Ae.)* mosquitoes in an urban transmission cycle.^13^ Though studies of wild-caught mosquito populations implicated the canopy-dwelling *Haemagogus (Hg.) janthinomys* mosquito as the primary vector during a major outbreak in Brazil,^9^ *Ae. aegypti* and *Ae. albopictus* have demonstrated the potential to transmit MAYV in laboratory settings.^14^

A recent epidemiological alert by the Pan American Health Association (PAHO) emphasized the need for increased awareness of and extended surveillance for MAYV in the Americas.^15^ Ideally, MAYV spillover and outbreak prevention in the Americas would be guided by granular maps of MAYV risk enabling targeted febrile surveillance and ecological surveillance efforts and better tailored risk communications to endemic populations and travelers. However, the precise areas of risk from MAYV throughout the Americas remain unclear due to limited data on MAYV occurrence^16^, underscoring a fundamental need for a more comprehensive and georeferenced dataset on MAYV occurrence in the Americas.

Here, we provide a critical update to the current state of knowledge on MAYV transmission risk across the Americas. We adopted a well-established machine-learning based disease mapping approach originally developed by ecologists to model species distributions but has since been successfully applied to several medically-relevant vector-borne pathogens^17–19^. These methods are particularly powerful for leveraging biological and ecological information underpinning a sylvatic disease system to generate biologically realistic and spatially explicit predictions when epidemiological data are still sparse. Many of these models rely on machine learning techniques including boosted regression trees (BRT)^20^ to develop a multivariate relationship between disease occurrence locations and relevant climatic or environmental covariates that impact disease transmission.

In order to develop a contemporary estimate of MAYV risk in the Americas, we applied a predictive mapping approach with three components: (1) scoring the total evidence supporting ongoing MAYV transmission within each country (i.e., evidence consensus scores); (2) modeling the likely distribution of MAYV suitability throughout the Americas with high predictive accuracy; (3) estimating the total population residing in areas with a high suitability for MAYV transmission. Compared to previous estimates, these updated datasets and analyses suggest that MAYV poses a substantial and possibly underestimated threat to the Americas.

## Methods

### Evidence Consensus

We used a well-established framework^21^ to generate an evidence consensus score for each country in the study region. This approach integrates several information sources to generate a score that characterizes the evidence for disease transmission. The data sources that we considered for MAYV evidence consensus scores included health organization status, date of most recent human occurrence, validity of diagnostic tests, recency of outbreaks or clinical cases, and recency of occurrence in non-human animals or arthropods. Scores ranged from 0 (“*No evidence of MAYV presence*”) to 21 (“*Complete evidence of MAYV presence*”). A more comprehensive description of the evidence consensus scoring process is presented in the Supplementary Materials (pp 3-5).

### Occurrence records

We collected and georeferenced MAYV occurrence records in humans, non-human animals, and arthropods^22, 23^. These records were extracted from peer-reviewed and grey literature sources. Occurrences with a high level of spatial precision were designated point locations, while less precise records (e.g., administrative units) were designated polygon locations. We recorded the coordinates of all point locations and polygon centroids as well as the uncertainty associated with each record. We included occurrence records with ≤75km of uncertainty in our current analysis. Overall, our dataset comprised 203 MAYV occurrence records in humans, animals, and arthropods. This was subsequently reduced to 170 records after we thinned the dataset to reduce spatial autocorrelation. Details on the assembly of the occurrence records are available in the Supplementary Materials (p 3).

### Description of covariates

We considered 10 ecologically relevant gridded variables (i.e., raster data) for inclusion in our model. These variables included various measures of topography, climate, land cover and vegetation that likely influence the MAYV transmission cycle and the distribution of MAYV risk throughout the region. (See Supplementary Materials for a description of the covariates and rationale for their inclusion in the model.) Values for each gridded covariate were extracted at each presence/pseudoabsence location and used in the modeling procedure described below. Climatic variables included rainfall, nighttime land surface temperature (LST Night), and daytime land surface temperature (LST Day) (Figures 1A – 1C). Measures of land cover and vegetation included two proportional land cover classes (evergreen broadleaf and urban/built-up), enhanced vegetation index (EVI), tasseled cap wetness (TCW) and tasseled cap brightness (TCB) (Figures 1D – 1H). Lastly, we included two topographic covariates: slope and elevation (Figures 1I and 1J). The covariate layers are presented as maps in Figure 1 and additional details on the covariates are presented in the Supplementary Materials (pp 5-6). We did not detect any collinearity between the covariates based on a Spearman correlation coefficient of 0.8, and thus included all 10 covariates in the final model.

**Figure 1.**
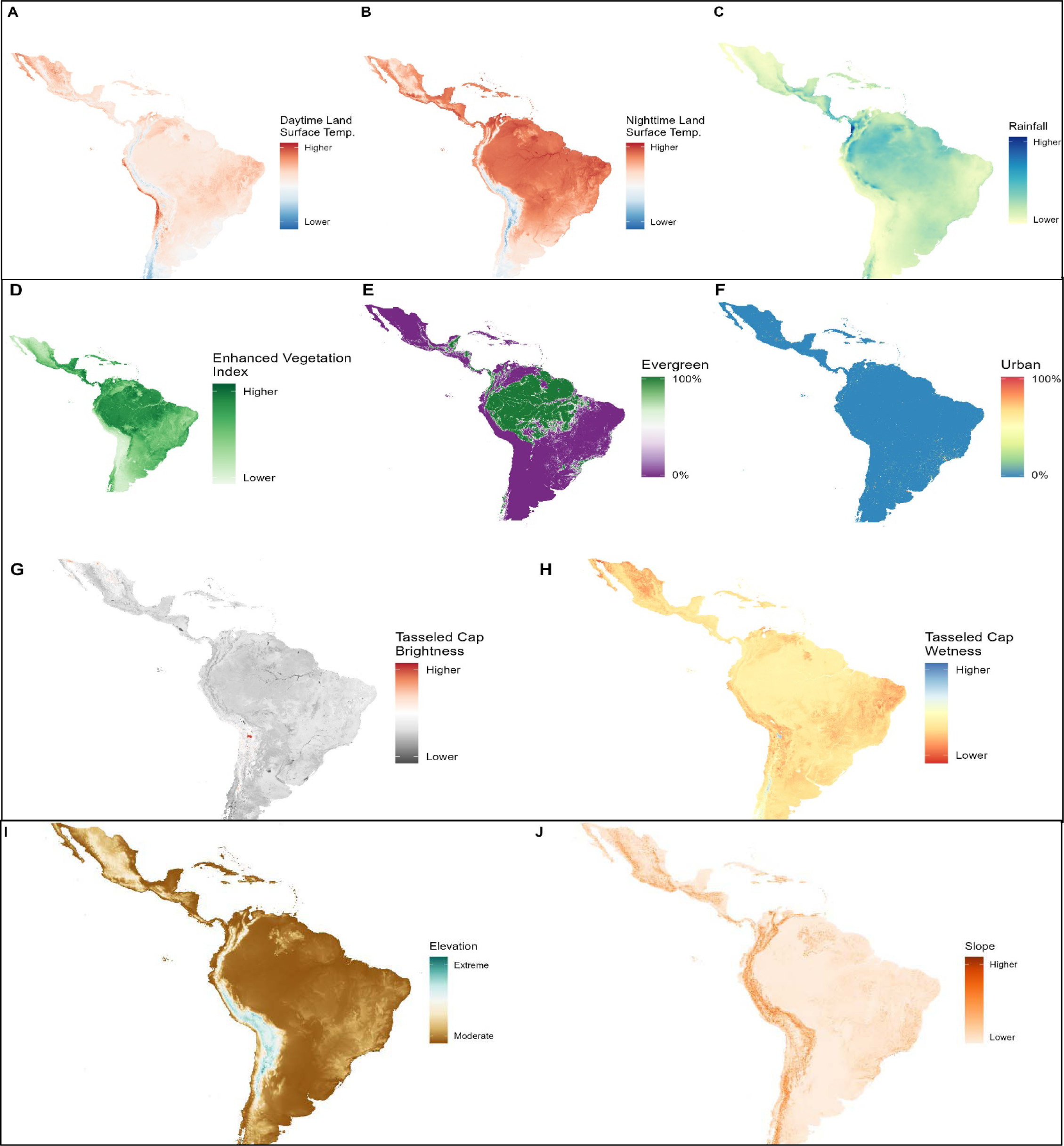
Covariates used to model the environmental suitability of MAYV. A. and B. Land surface temperature (LST) night and LST day, respectively; C. Rainfall; D. Enhanced vegetation index (EVI); E. Evergreen forest; F. Urban/built-up land cover; G. Tasseled cap brightness (TCB); H. Tasseled cap wetness (TCW); I. Elevation; J. Slope

### Mapping risk for MAYV occurrence

Although first developed by ecologists to model the potential distribution of plants and animals, ecological niche models (also known as species distribution models or environmental suitability models) are now common in the epidemiological literature to model human disease risk. A robust disease mapping framework has been established in the last decade to model the zoonotic niche of medically-relevant pathogens including dengue virus (DENV)^17^ and Zika virus (ZIKV)^18^, among others. We used BRT to model the environmental suitability for MAYV occurrence across our study region. BRT is a machine learning approach that uses regression or classification trees to partition the dataset and combines many simple models (i.e., boosting) to improve overall model accuracy.^20^ We fitted 100 BRT ‘sub-models’ to separate bootstraps of the dataset using the gbm.step procedure in the dismo R package.^24^ The predictive accuracy of the model was assessed using the area under the receiver operator characteristic curve (AUC), sensitivity, specificity, Kappa statistic, and percent correctly classified (PCC). Metrics were calculated for each sub-model using 10-fold cross validation. The final prediction map represents the mean MAYV suitability at each 5 x 5 km pixel across our ensemble of 100 models. We also generated a map of the model uncertainty, represented by the per-pixel standard deviation.

### Total population at potential risk

We estimated the total human population living in areas of high predicted MAYV suitability. We first transformed the mean prediction map into a binary risk map using a previously established protocol,^18^ whereby a suitability threshold value was chosen that encompassed 90% of the MAYV occurrence points. Each 5 x 5 km pixel was classified as 1 (i.e., suitable) if its predicted suitability exceeded the threshold value; otherwise, it was classified as 0 (i.e., not suitable). We then determined the total population residing in suitable areas by multiplying the population count within each grid cell by the binary suitability classification and summing these values across each country.

## Results

The map of evidence consensus is presented in Figure 2A and the evidence score for each country is presented by category in the Supplementary Materials (pp 11-16). Evidence consensus scores ranged from 0 (no evidence of MAYV transmission) to 19 (very high evidence of transmission). We recorded a very high evidence consensus score for Brazil and Venezuela, with scores of 19 and 16, respectively. Other countries with a high evidence consensus score included Peru with a score of 15, French Guiana with a score of 13, and Trinidad and Tobago, Colombia, and Bolivia (all with scores of 11). We recorded evidence consensus scores ranging from very low to moderate in all Central American and Caribbean countries. Among these countries with low to moderate risk, the highest evidence consensus scores were documented for Haiti with a score of 10, and Panama with a score of 9.

**Fig 2.**
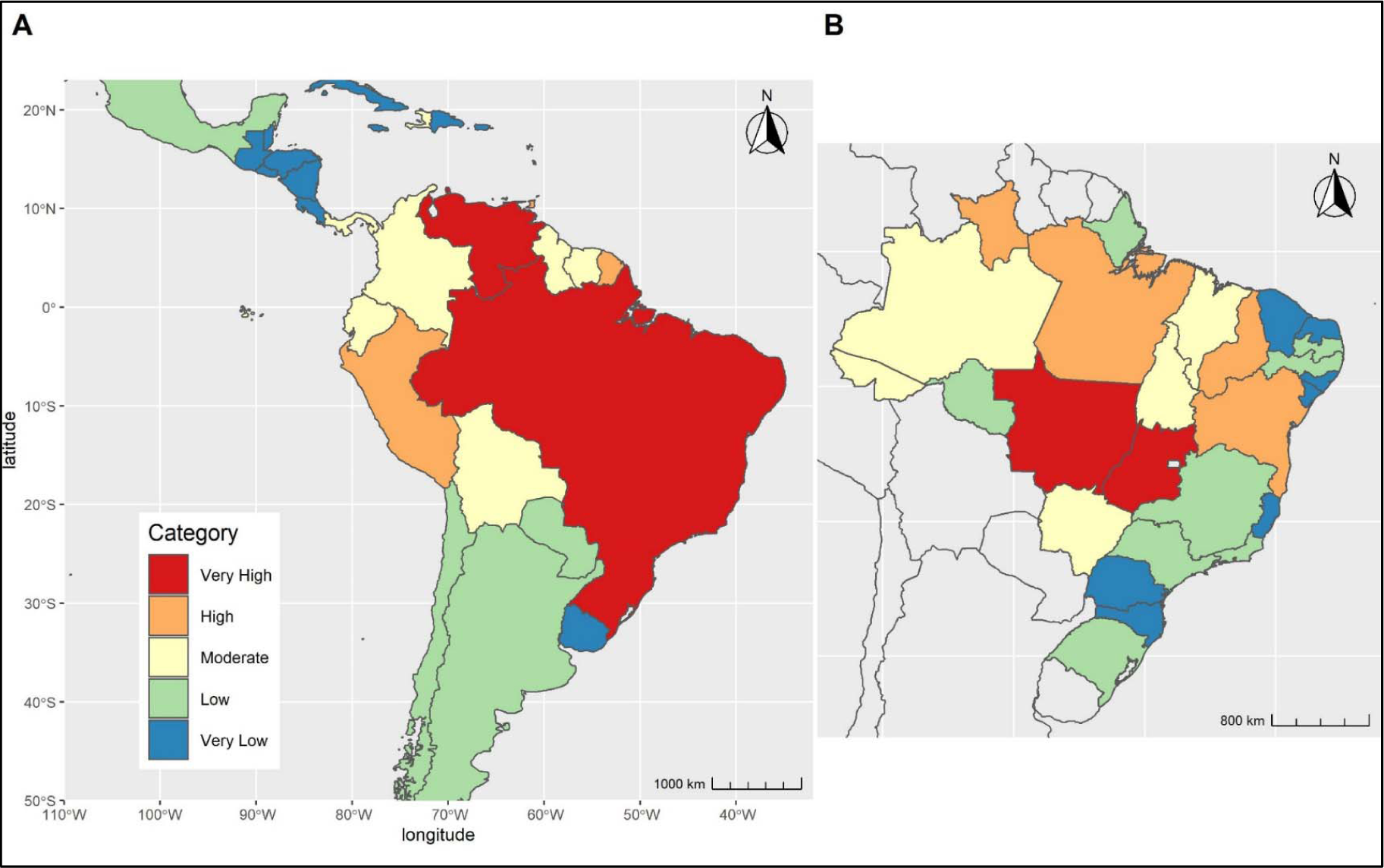
Evidence consensus scores. Evidence consensus is presented at the Admin0 level for all countries in the study (Fig. 2A) and at the Admin1 level (Fig. 2B) for Brazil. Scores are based on health organization status, date of most recent MAYV occurrence; validity of MAYV diagnostic test, recency of MAYV outbreaks or clinical cases, and recency of MAYV occurrence in animals or arthropods. Blue represents very low evidence consensus while red represents very high evidence consensus.

In order to provide more granular data throughout Brazil, we also calculated evidence scores by state (i.e., first-administrative division). These results are presented in Figure 2B. Because health organization status was not available for each state, we assigned a baseline score of one to each state, and then calculated the remaining categories according to the methods described above. Evidence of MAYV transmission was highest in the central Brazilian states of Mato Grosso and Goiás, both with very high scores of 16. High evidence of MAYV was also documented in five Northern and Central states, including Pará (Evidence Consensus= 14), Roraima (Evidence Consensus= 13), Bahia and Piauí (Evidence Consensus= 12 for both), and Mato Grosso do Sul (Evidence Consensus= 11).

Figure 3 displays the 203 MAYV occurrences (human, animal, and arthropod) that were used to construct our model. The occurrence locations fell in 13 countries, most frequently in Brazil (n=102), French Guiana (n=27) and Peru (n=22). MAYV occurrences were reported between the years 1954 and 2021, with the majority of cases (n=140) occurring since the year 2000. One hundred and sixty (79%) of the 203 occurrence locations were detected in humans while 43 (21%) were detected in non-human animals or arthropods. An alternative model using just the reported human occurrence was also constructed using the 160 human occurrence points. This model is presented in the Supplementary Materials along with a map showing the observed difference between the all-host and the human-only models (pp 8-9).

**Figure 3.**
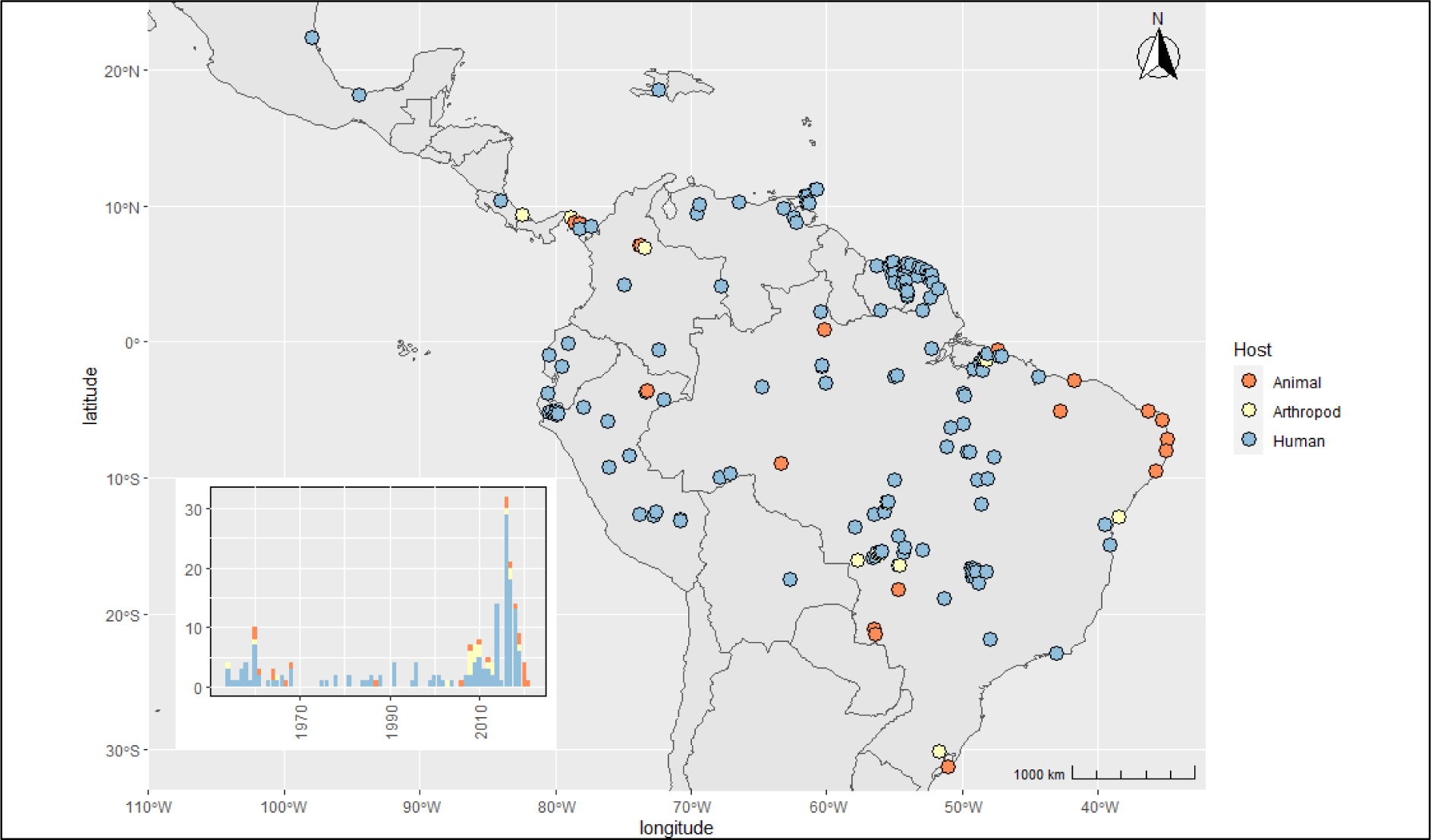
Geographic distribution and temporal trend of MAYV occurrence. The map shows the distribution of the 203 occurrence locations (before the spatial thinning procedure) that were used to construct the boosted regression tree (BRT) model. The color corresponds to the host type of each point (human, animal, or arthropod). The inset chart displays total occurrences that were reported in each year since the initial human case was detected in 1954.

The predicted distribution of MAYV environmental suitability is presented in Figure 4A. This risk map represents the average output across the 100 BRT sub-models. The map of model uncertainty (i.e., the per-pixel standard deviation across the 100 model runs) is presented in Figure 4B. High suitability for MAYV transmission was evident across the Amazon rainforest ecoregion in South America. The model predicted very high suitability for MAYV across a large portion of Central and Northern Brazil, especially the states of Amazonas, Acre, Pará, and Tocantins. High suitability was also predicted throughout French Guiana, Guyana, Suriname, and Trinidad and Tobago, as well as Southern portion of Colombia and Venezuela, and the north-eastern region of Peru and northern region of Bolivia.

**Fig 4.**
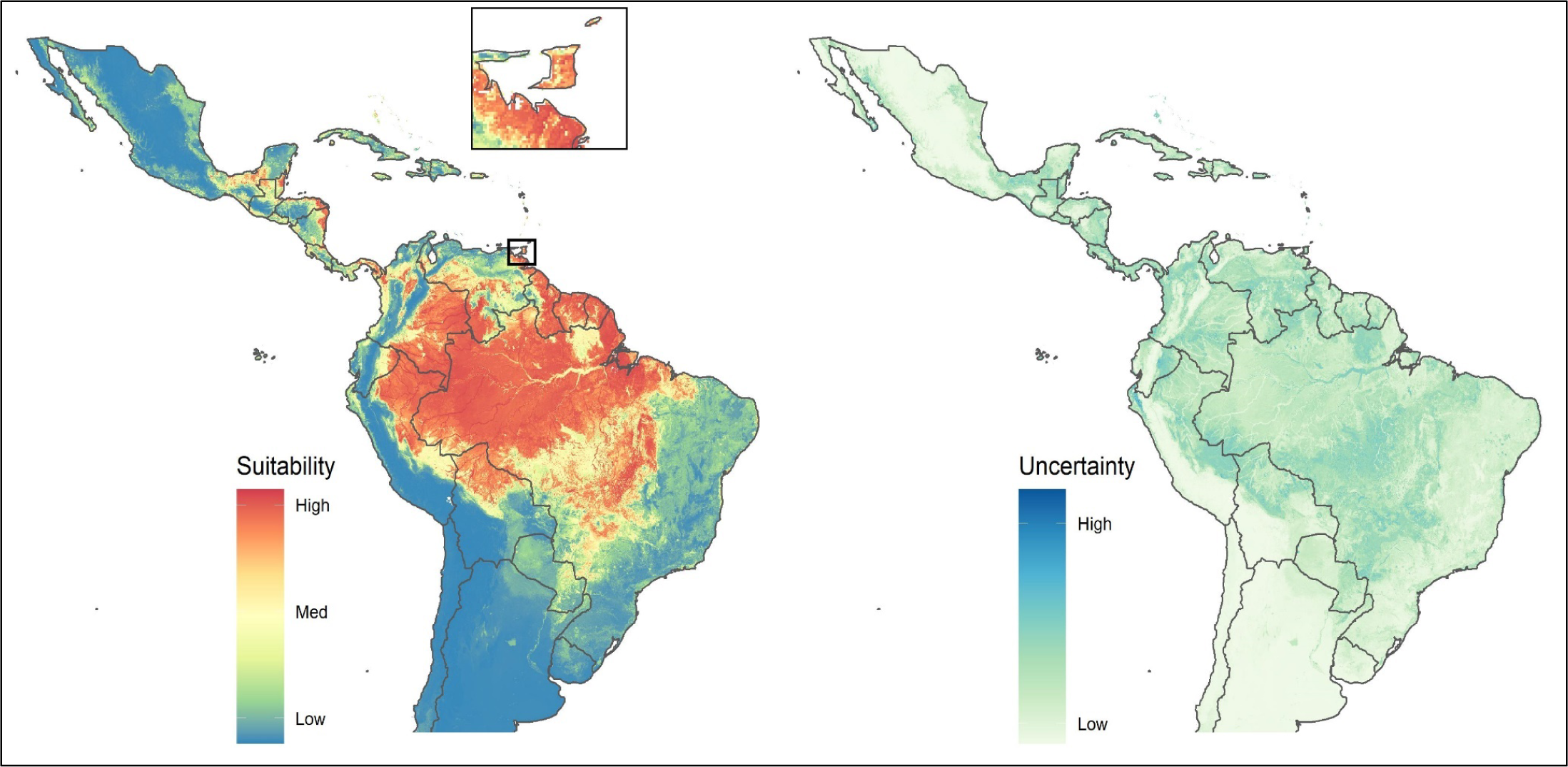
Map of environmental suitability and prediction uncertainty for MAYV occurrence. (A) Suitability ranged from blue (0 - no suitability) to red (1 - very high suitability). The inset map shows greater detail in Trinidad and Tobago. (B) The per-pixel standard deviation across the 100 sub-models is presented as a measure of the model’s uncertainty.

Certain regions of Central America with low or moderate evidence consensus scores were also found to be highly suitable for MAYV transmission, namely the Southern region of Panama and eastern coastal regions of Nicaragua, Honduras, and Belize. The results of the human-only model are presented in the Supplementary Figure 1. Overall, the results of this model were very similar to the all-host model, with only slight differences. The difference in predicted environmental suitability between the all-host model and human-only model is presented in Supplementary Figure 2.

Our models showed MAYV suitability to be especially influenced by climatic variables including nighttime LST (relative importance of 44·8%) and rainfall (relative importance of 23·6%) as well as EVI (relative importance of 8·1%). The partial dependence plots for nighttime LST and rainfall are presented in Figure 5 and partial dependence plots for the remaining variables are presented in the Supplementary Figure 3. The partial dependence plot for nighttime LST reveals a steep increase in MAYV suitability around 12℃ that peaks at ∼22℃. The plot for rainfall reveals a similarly steep increase starting at ∼80mm that peaks at ∼110mm and then plateaus, with only a minor peak at ∼375mm. After applying pairwise distance sampling to remove spatial sorting bias, the model demonstrated good predictive power with an AUC of 0·78 ± 0·007 standard error. Other statistics from the 10-fold cross-validation procedure included the following: PCC= 83%, sensitivity= 0·75, specificity= 0·91, and Kappa= 0·6.

**Fig 5.**
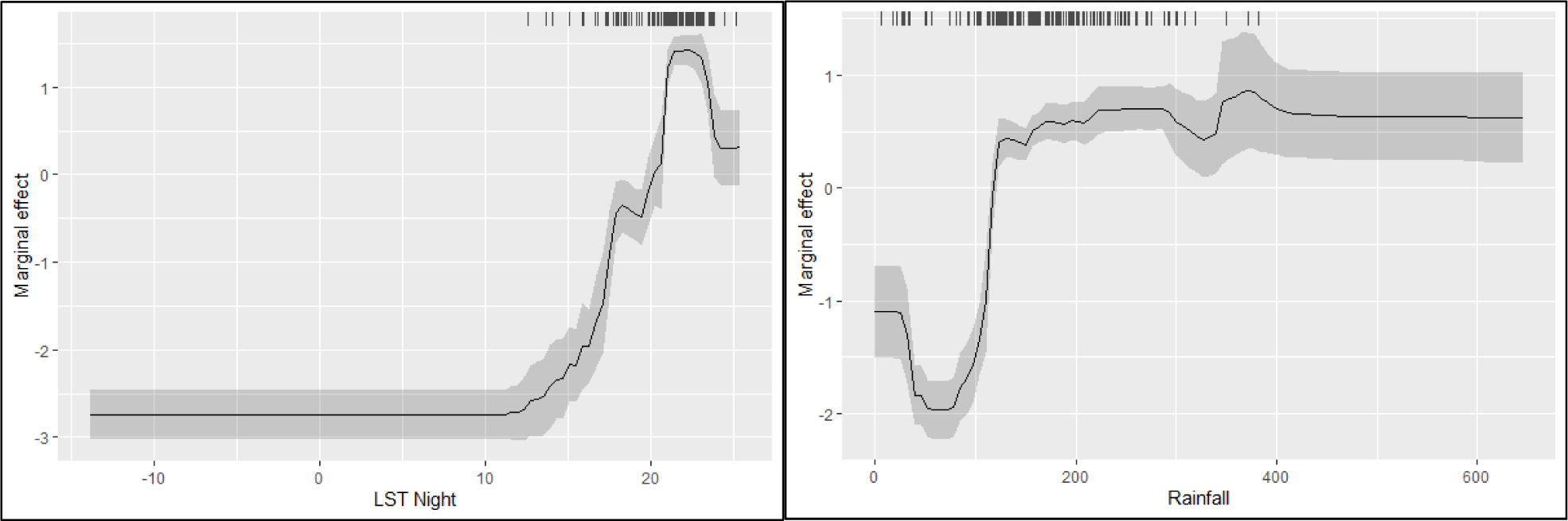
Partial dependence plots of the two most important variables. The solid black line represents average response over 100 sub-models and the gray region represents one standard deviation. Tick marks represent values of each variable at occurrence location.

We identified 0·488 as the threshold suitability value that encompassed 90% of the MAYV occurrence locations. We applied this conservative value to classify pixels as either suitable or unsuitable for MAYV transmission. Overall, we estimate that approximately 77 million people in the Americas live in areas that are potentially suitable for MAYV transmission. Countries with the greatest at-risk population include Brazil (43.4 million people), Colombia (6.9 million people), and Mexico (4.9 million people). The majority of the Brazilian population living in high-risk areas reside in the Amazon rainforest ecoregion.

**Table 1.** Total population living in areas potentially suitable for MAYV transmission (millions). Table 1 indicates the five countries in each region with the largest population in suitable transmission regions. The North/Central American region includes Caribbean Island nations.

| Country/Region | Pop. at risk (millions) |
| --- | --- |
| **South America** | **58.38** |
| Brazil | 43.37 |
| Colombia | 6.96 |
| Venezuela | 2.80 |
| Peru | 1.95 |
| Trinidad and Tobago | 0.90 |
| **North/Central America** | **18.33** |
| Mexico | 4.96 |
| Haiti | 3.09 |
| Dominican Republic | 2.52 |
| Panama | 2.30 |
| Guatemala | 1.21 |
| **Overall** | **76.71** |

## Discussion

In this paper we present an ensemble BRT model of MAYV environmental suitability in the Americas and an evidence consensus framework that integrates multiple information sources. In the absence of comprehensive epidemiological data and reliable and affordable methods for large scale arboviral diagnosis, distribution models can serve an important role in guiding arboviral surveillance (in humans and other hosts) and targeting vector control efforts. This is especially true in the case of MAYV given its nonspecific febrile presentation and the uncertainty surrounding its true distribution and non-human animal reservoirs. Our model provides important information regarding regions of Central and South America that are at highest risk of MAYV transmission allowing us to estimate the total population living in areas suitable for MAYV transmission. Furthermore, we are able to identify areas with high predicted MAYV suitability despite low or moderate evidence consensus.

Our model predicted the distribution of MAYV with relatively high accuracy, identifying several regions of high environmental suitability. This included large areas of North and Central Brazil (e.g., the states of Mato Grosso, Pará, and Goiás), French Guiana, Trinidad & Tobago, and Northern Peru, all areas with well-documented evidence of MAYV transmission.^25^ In addition, several regions with minimal or no published evidence of MAYV transmission were also found to be highly suitable for transmission, including the majority of Guyana as well as coastal regions of Belize, Nicaragua, and Honduras. This finding highlights the utility of distribution models in identifying areas that are particularly receptive to MAYV transmission that could be targeted for increased surveillance or vector control.

The wide predicted geographic distribution of MAYV underscores the need for increased surveillance and diagnostic capacity throughout the Americas. Our findings suggest that MAYV is likely vastly underreported and that co-occurring arboviral epidemics (e.g., DENV or CHIKV) may obfuscate the true MAYV disease burden. This is especially true in Brazil, where ∼43 million people reside in areas that are potentially suitable for MAYV transmission. An additional concern is the detection of MAYV in Haiti,^26^ where ∼3 million people are predicted to be at potential risk according to our model. The discovery of MAYV in Haiti has prompted additional questions about its potential vectors and the possibility of urban transmission, due to the lack of *Haemagogus* mosquitoes and non-human primates on the island. These questions require further entomological investigations in order to elucidate the vectorial capacity of urban mosquitoes and potential animal reservoirs other than non-human primates.

Our model has several important limitations that must be considered when interpreting the findings presented here. Georeferenced reports of disease occurrence are subject to sampling bias related to the accessibility of certain locations, availability of laboratory infrastructure, or the presence of a robust surveillance system that is able to detect arbovirus occurrence. Therefore, the presence locations used in our model are likely subject to sampling bias and do not reflect the true distribution of MAYV across the Americas. Following previously published modeling studies,^27^ we attempted to correct for the sampling bias in our dataset through the use of pseudoabsence points with a similar spatial bias as the presence points. However, it is likely that the model is still affected by some level of bias in the occurrence locations. Ideally, future MAYV distribution models will be updated when new occurrence locations are reported (as in Pigott et al.^28^ updates to the Ebola niche map), leading to a more accurate prediction of the true MAYV distribution.

Another limitation relates to the covariate set used to construct our model. Many aspects of the MAYV epidemiology remain poorly understood and current knowledge of MAYV ecology is lacking. We attempted to include covariates that likely have a strong influence on MAYV transmission, including climate, landcover, and vegetation indices, which impact vector ecology and therefore transmission risk. However, disease risk is influenced by other unknown or understudied variables including socioeconomic factors and the presence of non-human reservoir hosts. Although several non-human primate species appear to be important MAYV reservoirs,^9^ we opted not to include primate distribution in our model due to the lack of certainty regarding their precise role in the transmission cycle. For systems like YFV, where the role of particular primate species is better understood, the distributions of particular primate reservoir species may be incorporated directly into predictive models.^29^ As more research is conducted on MAYV ecology and additional animal reservoirs are identified, future MAYV distribution models should be updated to include primate distribution as a contributor to overall spillover risk. Our results here may assist in targeting and designing animal reservoir studies.

Our model contributes several crucial additions to the limited literature on MAYV distribution.^16^ By including a comprehensive compendium of MAYV occurrence points along with high-resolution, remotely sensed covariates at each point, the current ensemble model generates actionable predictions through its demonstrated high predictive accuracy^20^ across a wider geographic range than any previously published model. Furthermore, the evidence consensus scores provide an alternate method for estimating MAYV transmission, which complements the modeled predictions. These estimates thus inform where and when both human and non-human MAYV surveillance should be targeted. While it remains difficult to accurately estimate the costs of epidemic response^30^ our estimates of the total at-risk human population across multiple countries also make clear that the potential risk to human health is disproportionately high compared to investments in understanding the basic ecology and epidemiology of MAYV needed to mitigate the risk of future spillover transmission.

## Disclaimer

Material has been reviewed by the Walter Reed Army Institute of Research. There is no objection to its presentation and/or publication. The contents, views or opinions expressed in this publication are those of the authors and do not necessarily reflect official policy or position of Henry M. Jackson Foundation for the Advancement of Military Medicine, Inc., Uniformed Services University of the Health Sciences, Walter Reed Army Institute of Research, the Department of Defense (DoD), or Departments of the Army, Navy, or Air Force. Mention of trade names, commercial products, or organizations does not imply endorsement by the U.S. Government.

SP was supported by the National Institute of Allergy and Infectious Diseases, National Institutes of Health, https://www.niaid.nih.gov/, under Inter-Agency Agreement Y1-AI-5072, and the Defense Health Program, U.S. DoD, under award HU0001190002. AP and DP were financially supported by the Armed Forces Health Surveillance Division – Global Emerging Infections Surveillance (AFHSD-GEIS) award P0065_22_WR. The activities undertaken at the Walter Reed Biosystematics Unit were performed in part under a Memorandum of Understanding between the Walter Reed Army Institute of Research (WRAIR) and the Smithsonian Institution, with institutional support provided by both organizations. The funders had no role in study design, data collection and analysis, decision to publish, or preparation of the manuscript.

## Data Availability

All data produced in the present study are available upon reasonable request to the authors

## Appendices

### Occurrence records

We previously developed a georeferenced compendium of MAYV occurrence based on methods that have been established for other pathogens including dengue and leishmaniasis.^1–4^ MAYV occurrence among humans, non-human animals, and arthropods was compiled through a systematic review of the literature, including an evaluation of the quality of such evidence. These methods were described previously in greater detail.^5^ All occurrences were assigned to a point or polygon location, depending on the spatial resolution provided by the authors. Point data comprised precise locations with less than 5km of uncertainty (e.g., specific coordinates or a small town) while polygon data comprised larger areas or administrative units that exceeded 5km of uncertainty. The coordinates of point locations and polygon centroids were recorded in our database and used as the presence records in our current modeling study.

Presence points with ≤75km of uncertainty were included in our current analysis, although the majority of points had substantially less uncertainty. Following a previously published modeling study that accepted up to 65km of uncertainty,^6^ we chose to accept a greater level of uncertainty in our occurrence data in order to include more occurrence locations during the model development process. Due to the limited size of our occurrence dataset, we deemed that the extra information gained from each occurrence record outweighed any issues associated with the greater uncertainty of some occurrence records.

The final MAYV database contained 265 unique georeferences in 15 countries, published between 1954 and 2022. Two-hundred and three of these occurrence points met the ≤75km uncertainty threshold, and thus were eligible for inclusion in our model. We used the spThin package in the R statistical software to reduce clustering of presence records.^7^ A 5km distance threshold was applied to ensure that no more than one presence point occurred within each pixel of our covariate layers.

### Evidence Consensus

Collating published reports of MAYV is an important first step in clarifying its distribution. However, heterogeneous surveillance capacity across countries and incomplete or unclear reporting of epidemiological data may impact our ability to definitively say that MAYV is present in a certain location. An evidence consensus approach takes several information sources into account in order to score the total available evidence supporting the presence or absence of a disease in a given country. This approach has been used previously to provide a refined description of the spatial limits of several pathogens including dengue virus,^8^ leishmaniasis,^9^ podoconiosis,^10^ and Lassa fever.^11^ These studies considered multiple data sources to develop an evidence consensus score for disease presence or absence, including health organization status, peer reviewed evidence, case data, animal infection, economic status, and other supplementary evidence. We followed a similar procedure to generate a consensus score for each country in Latin America that quantifies the evidence supporting MAYV presence. This score ranged from 0 (“No evidence of MAYV presence”) to 21 (“Complete evidence of MAYV presence”) based on the categories described below.

### Health Organization Reports (max 3 points)

International health organizations have been used previously to support evidence of pathogen presence or absence in specific countries.^8^ We followed a similar procedure, using health reports from two sources: Pan American Health Organization (PAHO)/World Health Organization (WHO) and the Global Infectious Diseases and Epidemiology Online Network (GIDEON). The PAHO issues weekly epidemiological alerts to update the public on the occurrence of significant health events. Similarly, the WHO issues Disease Outbreak News (DONs) related to public health issues of international importance. We searched these PAHO/WHO bulletins for relevant alerts related to MAYV in a given country. Countries were assigned a score of 1 if WHO/PAHO had issued an epidemiological alert for MAYV in that country.

GIDEON is a web application that compiles relevant news on infectious disease outbreaks and designates countries as endemic/potentially endemic for each pathogen. If a country was listed as endemic/potentially endemic for MAYV in the GIDEON database, it was assigned a score of 1. If a country fulfilled both criteria (i.e., listed as endemic in GIDEON and a relevant PAHO/WHO health alert) it was assigned a score of 3.

### Peer-reviewed Evidence of Human Infection (max 6 points)

Based on methods proposed by Brady et al.,^8^ peer-reviewed evidence of human infection was scored based on the following two categories: contemporariness (3 for 2011-2020, 2 for 2000-2010, and 1 for 1999 and earlier) and diagnostic accuracy (3 for PCR, viral culture, or PRNT, 2 for serological methods only, 1 for presumed cases without diagnostic test or cases with unspecified diagnostic test). In the case of multiple MAYV reports in a given country, the highest scoring report was used. For example, if one study in Brazil reported serological evidence of MAYV transmission (score of 2) in 1990 (score of 1) and another study in Brazil reported MAYV viral culture (score of 3) in 2019 (score of 3), Brazil would receive a score of 6 for this category. We also considered returning traveler reports for this category if the case was definitively linked to the country of travel. These reports are useful for establishing evidence of pathogen presence because diagnosis is often pursued rigorously for travelers upon returning to their country of origin.^8^

### Outbreaks and Clinical Cases (max 6 points)

Reported outbreaks of MAYV or clinical cases that were detected using PCR were scored according to total case numbers and contemporariness. Previous studies have used only the occurrence of reported outbreaks (with no consideration to clinical cases) to assign a score for this category.^8^ However, because only a limited number of MAYV outbreaks have occurred and because diagnostic techniques have been inconsistent across these outbreaks, we also considered clinical cases diagnosed by PCR or viral culture that were not necessarily considered to be an outbreak. The scoring system for this category was adapted based on methods used by Mylne et al.,^11^ where higher scores were assigned to contemporary reports with 20 or more cases. In order to calculate a score for each country, we summed the cases detected across multiple studies within a single time period. For example, if two separate reports from Peru each detected 10 cases of MAYV using PCR between 2011 and 2020, we summed the reported cases (20 total) and assigned Peru 6 points for this category. However, cases from multiple reports were not summed across different time periods.

A lack of MAYV case reports in a given country is not necessarily indicative of a lack of virus transmission. It is likely that MAYV cases may go undetected due to insufficient surveillance or diagnostic capacity. We attempted to account for this uncertainty using healthcare expenditure (HE) as a proxy of a country’s capacity for detecting MAYV occurrence. If no outbreaks or clinical cases were reported in a given country, we used the current HE per capita from the World Health Organization 2017 dataset. Total HE for each country was designated as low (HE < $100), medium ($100 ≤ HE < $500), or high (HE ≥ $500) according to methods that were previously described.^8^ We also considered a country’s proximity to other countries that have reported outbreaks or clinical cases diagnosed by PCR.^11^ Adjacency to countries with outbreaks or clinical cases was combined with HE to assign a score. The highest score was assigned to countries with MAYV outbreaks/clinical cases in two or more neighboring countries and a low HE.

### Animal and Arthropod Data (max 6 points)

Detection of MAYV occurrence in non-human animal or arthropod species provides additional evidence of MAYV presence. This may be indicative of the potential for disease spillover into the human population. Previous studies have considered the presence of competent vector species when calculating the evidence consensus score.^12, 13^ However, because of uncertainties regarding the role of various mosquitoes in the MAYV transmission cycle (e.g., the possible role of *Aedes aegypti* in urban transmission^14^), we considered reports of any wild-caught arthropods that were identified as MAYV positive. Similarly, we assigned a separate score based on reports of infection in potential animal reservoirs. The highest scores for both animal and arthropod MAYV positivity were assigned to more contemporary studies. In the case of multiple MAYV reports in a given country, the highest scoring report was used. All data on MAYV positivity in non-human animals and arthropods was compiled in a systematic review that was previously described.^5^

### Explanatory covariates

We considered several variables for inclusion in our models based on both previous studies of and hypotheses about MAYV ecology. Rainfall data and remotely sensed variables from the MODIS platform were provided by the Malaria Atlas Project (https://malariaatlas.org/) after a gap-filling algorithm was used to account for cloud cover.^15^ The variables were transformed to ensure matching spatial resolution of 2·5 arc-minutes (∼5km) and matching extent. The variables were transformed to ensure that spatial resolution, extent, and boundaries were identical before modeling. Several variables described here were derived from NASA’s Moderate Resolution Imaging Spectroradiometer (MODIS) remote sensing platform.^16^

Temperature and rainfall play an important role in vector abundance and activity.^17^ Entomological surveys have demonstrated an association between *Hg. janthinomys* abundance and temperature^18, 19^ and Alencar et al. reported that the mosquito’s presence was correlated with high temperatures ranging from 24℃–30℃.^20^ Several studies have also demonstrated that large diurnal temperature range can impact larval development time, adult survival, and reproductive output in *Aedes* and *Anopheles* populations^21–23^ and an ecological niche model demonstrated that mean diurnal range was one of the most important predictors of *Hg. janthinomys* distribution.^24^ Humidity and rainfall have also been shown to impact the density of adult *Hg. janthinomys* mosquito populations^18, 25–28^ and *Hg. janthinomys* biting activity was shown to peak during intense rainfall in January.^26^ Due to the impact of temperature and rainfall on vector abundance and vectorial capacity, we included three climate variables in our model, namely night-time and daytime land surface temperature (LST) and cumulative rainfall. LST Night and LST Day are remotely sensed variables from the NASA MODIS MOD11A2 satellite.^29^ Annual LST Day and LST Night raster layers spanning the years 2000–2020 were used to calculate a single layer representing the mean values over this time period. We also used rainfall data from the Climate Hazards Group InfraRed Precipitation with Station (CHIRPS),^30^ a quasi-global data set that incorporates satellite imagery at 0·05° resolution and meteorological station data to construct gridded rainfall estimates.

In addition, *Hg. janthinomys* mosquitoes thrive in arboreal habitats (e.g., tropical rainforests) and oviposit in water-filled natural plant cavities (e.g., tree holes or broken bamboo).^31^ Adult mosquitoes have predominantly been found in the forest canopy at heights of 16m and 30m.^18^ Therefore, the density of vegetation canopy and moisture supply may influence *Hg. janthinomys* abundance in a given area. To account for these factors, we included three covariates related to vegetation and surface moisture: enhanced vegetation index (EVI), tasseled cap wetness (TCW) and tasseled cap brightness (TCB). The EVI is a measure of vegetation canopy greenness displayed at a 500m spatial resolution^32^ and is derived from the MODIS MCD43B4 product.^33^ The EVI has been used as a covariate in previous niche models of arboviruses including Yellow Fever,^34^ chikungunya virus,^12^ and Zika virus.^35^ TCW and TCB, measures of surface moisture that are used to assess land cover change, were also generated from the MODIS MCD43B4 product.^36^

Previous outbreaks of MAYV have occurred in towns close to the rainforest or jungle outposts in close proximity to the forest edge.^37–39^ MAYV most likely circulates in a sylvatic cycle involving canopy-dwelling mosquitoes and non-human primates, with occasional spill-overs into humans living close to the forest.^40^ Entomological surveys have demonstrated that *Hg. janthinomys* are predominantly found in forest canopies at heights of 16m and 30m.^18^ Due to the strong impact of land cover on the probability of MAYV occurrence in a given area, we included two land cover covariates, namely evergreen broadleaf forest and urban/built-up, from the MODIS MOD13Q1 product.^41, 42^ These covariates represent the proportion of each raster grid cell (ranging from 0-100) that is covered by the land cover class in question, whereby a value of zero represents the absence of land cover and 100 represents complete coverage. Lastly, we included slope and elevation covariates that we accessed from the US Geological Survey’s Global Multi-resolution Terrain Elevation Data (GMTED).^43^

### Covariate selection

We implemented a data-driven variable selection process in the R package SDMtune to identify variables for inclusion in our models. This process involves removal of highly correlated variables based on an algorithm that first ranks the variables by permutation importance and evaluates the correlation between the most important variable and the remaining variables. A leave-one-out Jack-knife test is then used to remove the variable that has the smallest impact on model performance according to the AUC. However, no variables met the threshold for exclusion based on a Spearman correlation coefficient of 0·8. Therefore, all variables were included in the analysis.

### Modelling approach

We used boosted regression trees (BRT) to model the environmental suitability for MAYV occurrence. BRT is a machine learning approach that has been used extensively to develop risk maps of vector-borne pathogens.^12, 13, 35, 44, 45^ This algorithm uses regression or classification trees to partition the dataset using recursive binary splits. It also incorporates boosting into the model-building process, a procedure that combines many simple models to improve overall model accuracy. The boosting algorithm is an iterative process that fits many small trees sequentially, building on previously fitted trees to improve model performance.^46^ This process incorporates a level of stochasticity by randomly selecting a subset of the data to fit each tree, thereby reducing the model variance.^46^ BRT have several advantages including their ability to fit complex nonlinear relationships, handle missing data, and to accommodate many different types of covariates without any need for data transformation.^46^

One of the most important aspects of modeling species distributions with presence-only data is the selection of pseudo-absence points that represent the range of environmental conditions where the species or pathogen was not detected. Random selection of pseudo-absence points may not be appropriate if the presence locations are spatially biased.^47^ In most cases, the detection of disease presence locations may be subject to sampling bias if some locations are more likely to be surveyed than others (e.g., locations that are closer to roads).^48^ Therefore, pseudo-absence points should be selected with a similar level of bias as the presence points to ensure that background and presence locations are biased in the same manner.^48^ Following the methods of previous modeling studies,^11, 49–51^ we selected 10,000 background points from the study region, biased towards more populous areas. Therefore, population density was used as a proxy for sampling bias. Pseudoabsence points were selected using the 2° method proposed by Barbet-Massin (2012),^52^ whereby each pseudo-absence point was at least 2° away from a presence location. In order to improve the model’s discrimination capacity, the pseudoabsence points were down-weighted to ensure that the weighted sum of presence records equaled the sum of weighted background points.^52^

We subsequently fitted 100 BRT ‘sub-models’ to separate bootstraps of the dataset. The bootstrapped datasets were chosen with replacement, subject to the condition that a minimum of 25 presence and 25 pseudo-absence points were selected. This bootstrapping procedure allowed us to quantify the uncertainty across models and to increase the model’s robustness.^53^ Each sub-model was fit in R using the gbm.step procedure in the dismo package.^54^ This function uses cross-validation to identify the optimal number of trees for each sub-model to improve predictive capacity. The remaining BRT hyperparameters were held at their default values (tree complexity = 4, learning rate = 0·005, bag fraction = 0·75, cross-validation folds = 10, step size = 10). The final prediction map represents the mean MAYV suitability of each 5 x 5 km pixel across our ensemble of 100 models. We also generated a map of the model uncertainty, represented by per-pixel standard deviation. In order to avoid extrapolating the model to regions far outside of the MAYV niche, the model predictions were restricted to the Americas.

Two models were constructed. The model presented in the main text (“all-host”) included all human, arthropod, and non-human animal occurrence data (203 occurrence points overall and 170 points after the spatial thinning procedure). An additional model presented in the Supplementary materials (“human-only”) included only human occurrence data (160 occurrence points overall and 143 points after the spatial thinning procedure). Similar to a previous model of Marburg virus risk,^51^ we opted to use all occurrence points in the primary model to leverage all available data. We were also interested in exploring how the model predictions might change with the inclusion of multiple host types. We compared the output of the two models by subtracting the raster pixel values, allowing us to visualize the difference in predicted suitability between the human-only model and the all-host model. The all-host model is presented in the results below while the human-only model and comparison of model output is presented in Supplementary Materials.

### Model evaluation

The predictive accuracy of the model was assessed using several performance metrics including the area under the receiver operator characteristic curve (AUC), sensitivity, specificity, Kappa statistic, and percent correctly classified (PCC). Metrics were calculated for each sub-model using 10-fold cross validation. The cross-validation procedure involved randomly splitting each bootstrapped dataset into 10 folds with approximately the same number of presence and absence records in each fold. The model was subsequently trained on nine of the folds and the withheld fold was used to evaluate the model performance. The performance metrics for each sub-model represent the mean values across the 10 folds. These values were then averaged across each of the 100 sub-models to generate an estimate of overall model performance.

---

**Supplementary Figure 1.**
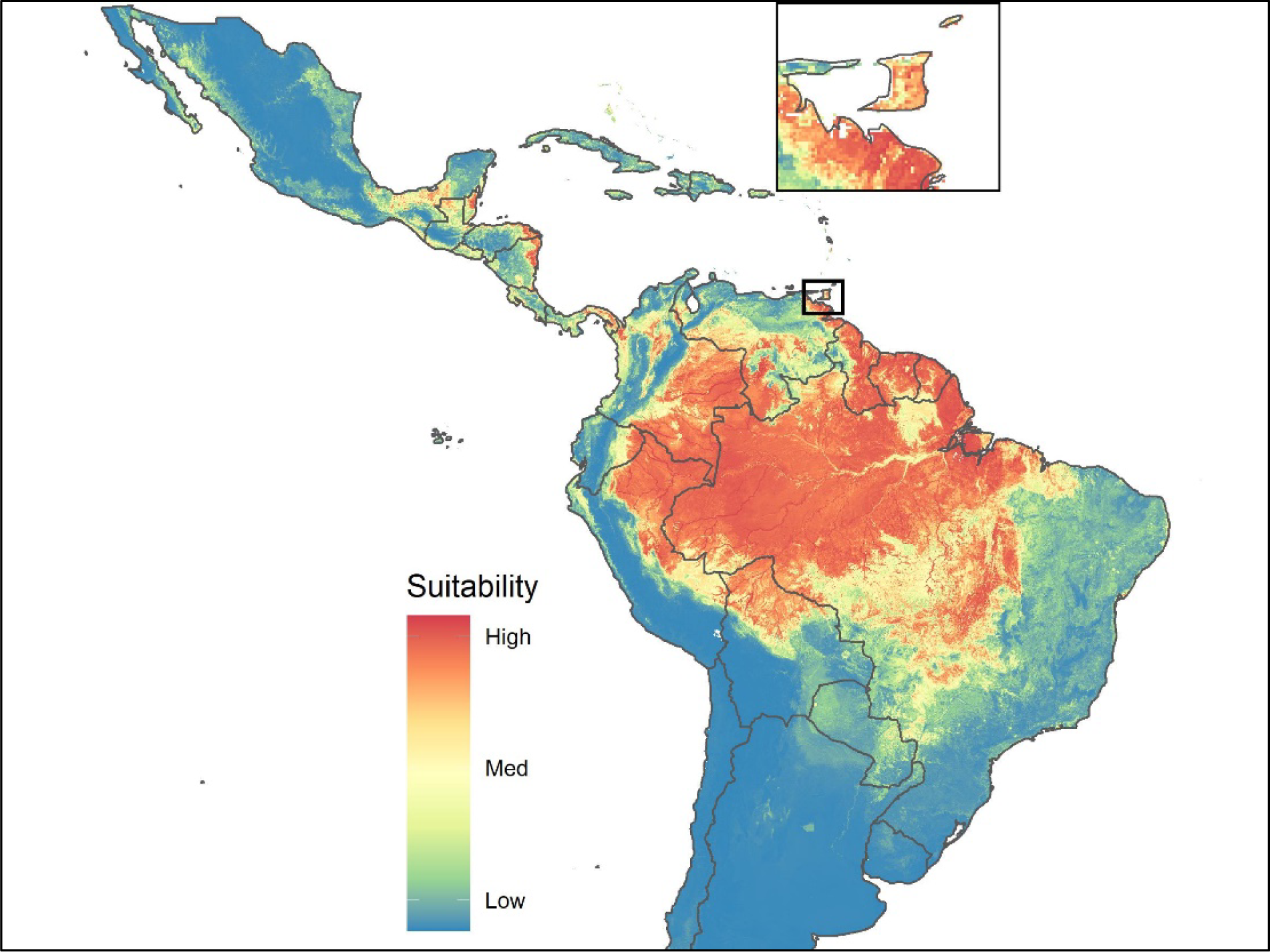
Map of environmental suitability MAYV occurrence using human-only data. Suitability ranged from blue (0- no suitability) to red (1- very high suitability). The inset map shows greater detail in Trinidad and Tobago.

**Supplementary Figure 2.**
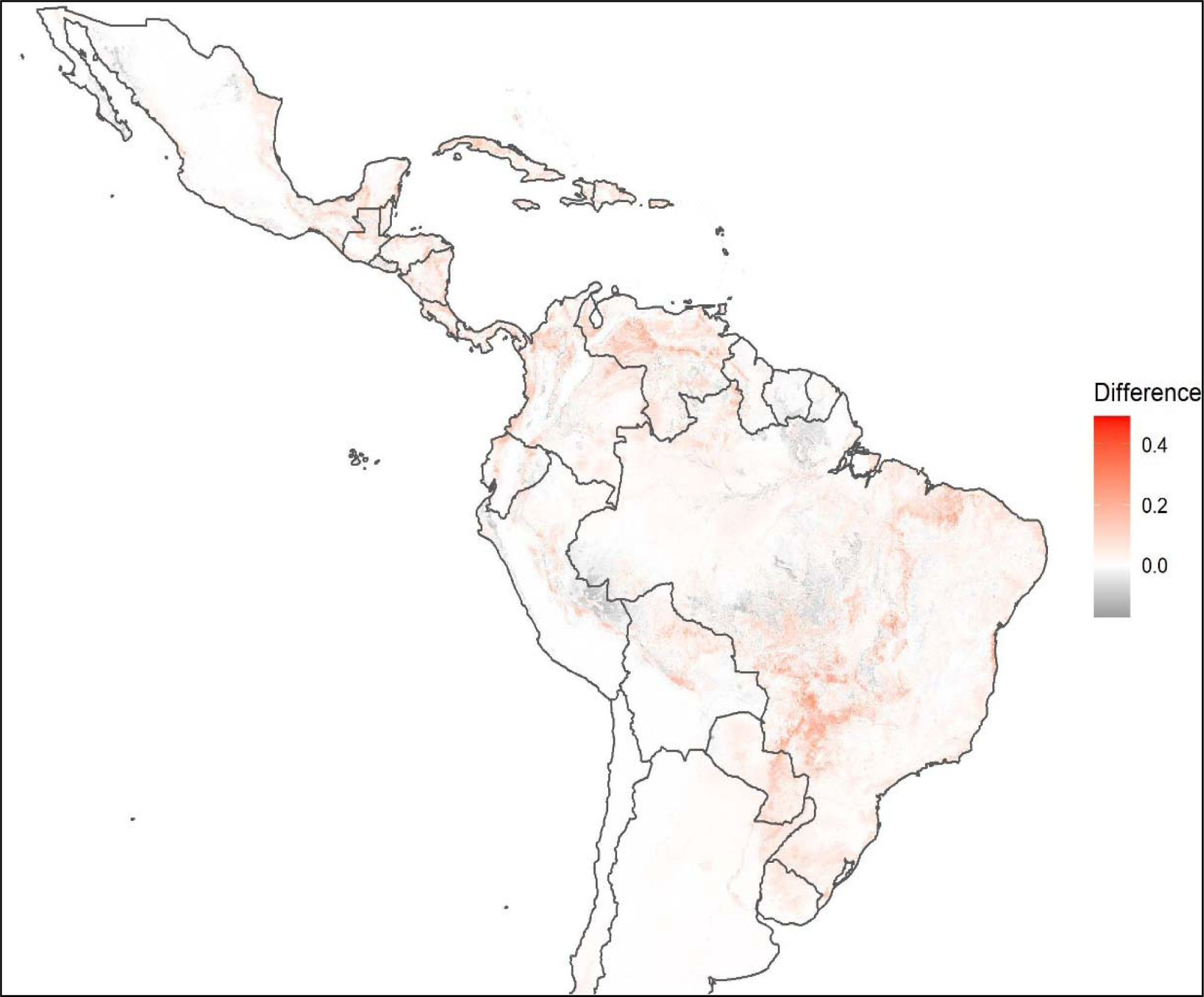
Difference in predicted environmental suitability between the all-host model and human-only model. This map was creating by subtracting the pixel-wise probability values in the all-host suitability model from the values in the human-only model. Red represents regions where the all-host model predicted higher suitability while black represents areas where the human-only model predicted higher suitability. White pixels represent agreement between the two models.

**Supplementary Figure 3.**
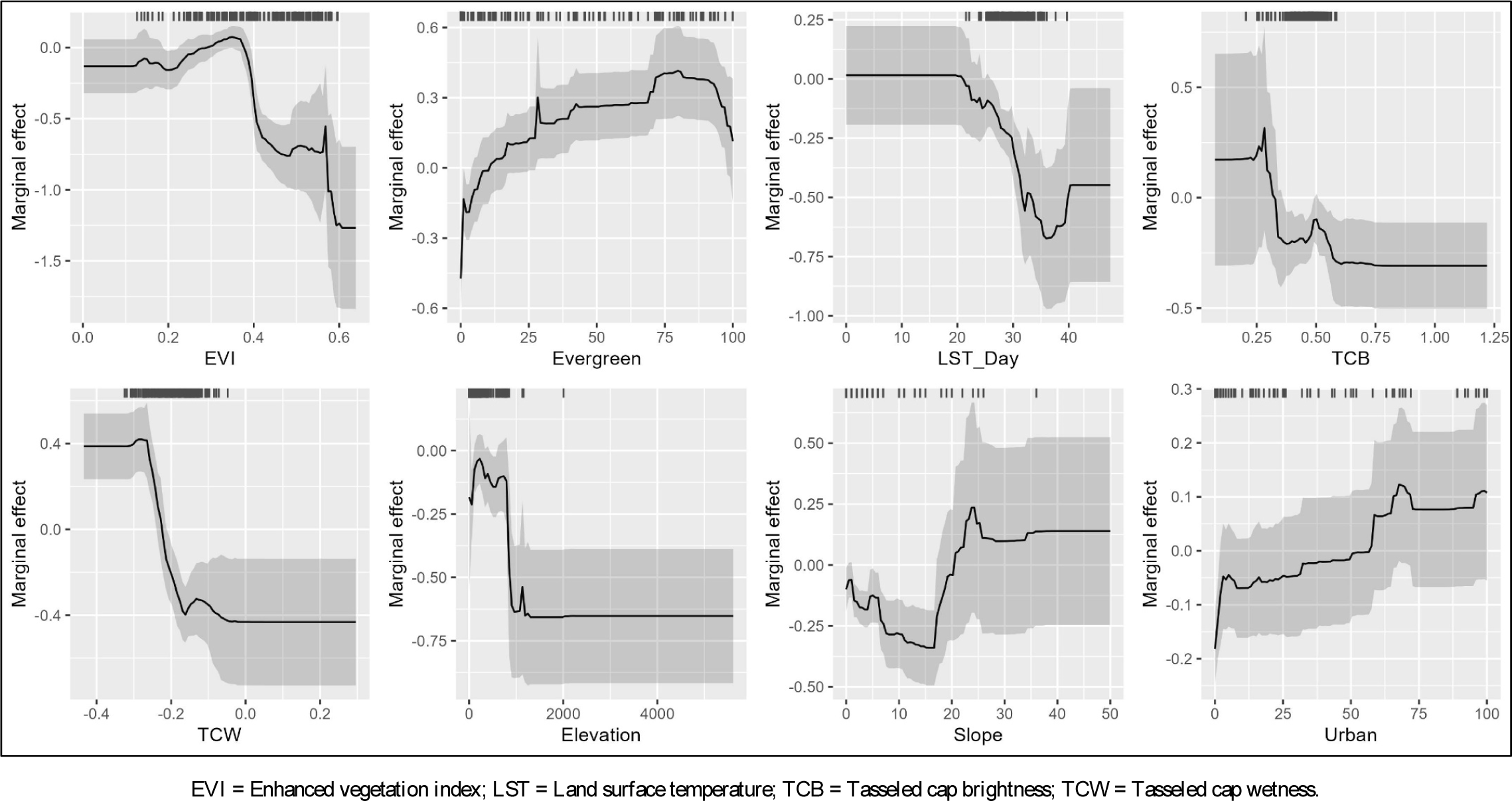
Partial dependence plots of the two eight additional variables. The solid black line represents average response over 100 sub-models and the gray region represents one standard deviation. Tick marks represent values of each variable at occurrence location. Partial dependence plots for the two most important variables are presented in the main text (Figure 5).

**Supplementary Table 1.** Evidence Categories and Possible Scores

| **Evidence Category** | **Score** |
| --- | --- |
| Health organization status |  |
| Both GIDEON and PAHO/WHO | 3 |
| Either GIDEON or PAHO/WHO | 1 |
| Peer reviewed evidence |  |
| Date of MAYV human occurrence |  |
| 2011-2020 | 3 |
| 2000-2010 | 2 |
| 1999 and earlier | 1 |
| Diagnostic procedure |  |
| PCR or viral culture or PRNT | 3 |
| Serological methods (not including PRNT) | 2 |
| Presumptive diagnosis or not specified | 1 |
| Outbreaks and clinical cases |  |
| 20+ cases from 2011-2020 | 6 |
| 20+ cases from 2000-2010 | 5 |
| 20+ cases 1999 and earlier | 4 |
| <20 cases from 2011-2020 | 3 |
| <20 cases from 2000-2010 | 2 |
| <20 cases 1999 and earlier | 1 |
| If no case data: health expenditure in 2017 and adjacency |  |
| HE <100 USD and 2 or more neighbors | 4 |
| 100 USD ≤ HE <500 USD and 2 or more neighbors | 3 |
| HE <100 USD and 1 neighbor | 2 |
| 100 USD ≤ HE <500 USD and 1 neighbor | 1 |
| Animal data |  |
| Infected animal from 2011-2020 | 3 |
| Infected animal from 2000-2010 | 2 |
| Infected animal 1999 and earlier | 1 |
| Arthropod data |  |
| Positive arthropod from 2011-2020 | 3 |
| Positive arthropod from 2000-2010 | 2 |
| Positive arthropod 1999 and earlier | 1 |
| **Possible Evidence Consensus Score Categories (Maximum Possible Score: 21)** |  |
| **Very high evidence of MAYV presence** | **16-21** |
| **High evidence of MAYV presence** | **11-15** |
| **Moderate evidence of MAYV presence** | **6-10** |
| **Little to no evidence of MAYV presence** | **0-5** |

**Supplementary Table 2:** Evidence consensus score by country

| Country | Health Org. Status | Date of Human Occurrence | Diagnostic Procedure | Outbreaks and Clinical Cases | Health Expenditure and Adjacency | Animal Data | Arthropod Data | Total Score |
| --- | --- | --- | --- | --- | --- | --- | --- | --- |
| **Brazil** | **1** | **3** | **3** | **6** | **N/A** | **3** | **3** | **19- Very high** |
|  | GIDEON only | Most recent occurrence: 2021<sup>55</sup> | RT-PCR<sup>56</sup> | 26 clinical cases in 2017-18<sup>56</sup> |  | Detected in horses in 2016<sup>57</sup> | Detected in mosquitoes 2018-19<sup>58</sup> |  |
| **Venezuela** | **3** | **3** | **3** | **6** | **N/A** | **1** | **0** | **16- Very high** |
|  | GIDEON and PAHO alert<sup>59</sup> | Most recent occurrence: 2016<sup>60</sup> | Isolation<sup>60</sup> | 77 clinical cases during outbreak in 2010<sup>61</sup> |  | Detected in sentinel hamster in 1999<sup>62</sup> | None detected |  |
| **Peru** | **1** | **3** | **3** | **6** | **N/A** | **2** | **0** | **15- High** |
|  | GIDEON only | Most recent occurrence: 2019<sup>63</sup> | RT-PCR and isolation<sup>64</sup> | 86 clinical cases in 2016<sup>65</sup> |  | Detected in various animals in 2008<sup>66</sup> | None detected |  |
| **French Guiana** | **3** | **3** | **3** | **3** | **N/A** | **1** | **0** | **13- High** |
|  | GIDEON and PAHO alert<sup>67</sup> | Most recent occurrence: 2020<sup>67</sup> | RT-PCR<sup>67</sup> | 13 clinical cases in 2020<sup>67</sup> |  | Detected in various animals in 1994-95<sup>68</sup> | None detected |  |
| **Trinidad & Tobago** | **1** | **3** | **3** | **3** | **N/A** | **0** | **3** | **13- High** |
|  | GIDEON only | Most recent occurrence: 2014<sup>69</sup> | RT-PCR<sup>69</sup> | 9 clinical cases in 2014<sup>69</sup> |  | None detected | Identified by metatranscriptomics in 2018<sup>70</sup> |  |
| **Bolivia** | **1** | **2** | **3** | **5** | **N/A** | **0** | **0** | **11- High** |
|  | GIDEON only | Most recent occurrence: 2007<sup>71</sup> | RT-PCR and isolation<sup>71</sup> | 23 clinical cases from 2000-07<sup>71</sup> |  | None detected | None detected |  |
| **Colombia** | **1** | **1** | **3** | **0** | **4** | **1** | **1** | **11- High** |
| | GIDEON only | Most recent occurrence: 1966<sup>72</sup> | NT<sup>73</sup> | No clinical cases | Per capita CHE was \$513; adjacent to Brazil, Venezuela, Peru | Detected in primates in 1957<sup>73</sup> | Isolated from mosquito in 1958-60<sup>74</sup> | |
| **Haiti** | **1** | **3** | **3** | **3** | **N/A** | **0** | **0** | **10- Moderate** |
|  | GIDEON only | Most recent occurrence: 2015<sup>75</sup> | RT-PCR<sup>75</sup> | 5 clinical cases 2014-15<sup>75</sup> |  | None detected | None detected |  |
| **Ecuador** | **1** | **3** | **3** | **2** | **N/A** | **0** | **0** | **9- Moderate** |
|  | GIDEON only | Most recent occurrence: 2019 <sup>76</sup> | Isolation, RT-PCR, or IgM seroconversion (not specified) <sup>71</sup> | 1 clinical case 2000-07 <sup>71</sup> |  | None detected | None detected |  |
| **Guyana** | **1** | **1** | **2** | **0** | **5** | **0** | **0** | **9- Moderate** |
| | GIDEON only | Most recent occurrence: 1955 <sup>77</sup> | Serology (not specified) <sup>77</sup> | No clinical cases | Per capita CHE was \$296; adjacent to Brazil and Venezuela | None detected | None detected | |
| **Panama** | **1** | **3** | **3** | **0** | **0** | **1** | **1** | **9- Moderate** |
|  | GIDEON only | Most recent occurrence: 2017 <sup>78</sup> | NT <sup>78</sup> | No clinical cases | No neighbors | Detected in various animals in 1974-76 <sup>79</sup> | Isolated from mosquito in 1972-79 <sup>80</sup> |  |
| **Suriname** | **1** | **1** | **3** | **0** | **3** | **0** | **0** | **8- Moderate** |
| | GIDEON only | Most recent occurrence: 1964 <sup>81</sup> | Isolation <sup>82</sup> | No clinical cases | Per capita CHE was \$474; adjacent to Brazil and French Guiana | None detected | None detected | |
| **Mexico** | **1** | **2** | **2** | **0** | **0** | **0** | **0** | **5- Low** |
|  | GIDEON only | Most recent occurrence: 2001 <sup>83</sup> | IgM ELISA <sup>83</sup> | No clinical cases | No neighbors | None detected | None detected |  |
| **Paraguay** | **0** | **0** | **0** | **0** | **5** | **0** | **0** | **5- Low** |
| | Neither | No cases | No cases | No clinical cases | Per capita CHE was \$400; adjacent to Brazil and Bolivia | None detected | None detected | |
| **Argentina** | **0** | **0** | **0** | **0** | **4** | **0** | **0** | **4- Low** |
| | Neither | No cases | No cases | No clinical cases | Per capita CHE was \$1128; adjacent to Brazil and Bolivia | None detected | None detected | |
| **Chile** | **0** | **0** | **0** | **0** | **4** | **0** | **0** | **4- Low** |
| | Neither | No cases | No cases | No clinical cases | Per capita CHE was \$1456; adjacent to Bolivia and Peru | None detected | None detected | |
| **Costa Rica** | **0** | **1** | **2** | **0** | **0** | **0** | **0** | **3- Low** |
|  | Neither | Most recent occurrence: 1968 <sup>84</sup> | HI test <sup>84</sup> | No clinical cases | No neighbors | None detected | None detected |  |
| **Uruguay** | **0** | **0** | **0** | **0** | **1** | **0** | **0** | **1- Low** |
| | Neither | No cases | No cases | No clinical cases | Per capita CHE was \$1590; adjacent to Brazil | None detected | None detected | |
| **Dominican Republic** | **0** | **0** | **0** | **0** | **1** | **0** | **0** | **1- Low** |
| | Neither | No cases | No cases | No clinical cases | Per capita CHE was \$462; adjacent to Haiti | None detected | None detected | |
| **Belize** | **0** | **0** | **0** | **0** | **0** | **0** | **0** | **0- Low** |
|  | Neither | No cases | No cases | No clinical cases | No neighbors | None detected | None detected |  |
| **Guatemala** | **0** | **0** | **0** | **0** | **0** | **0** | **0** | **0- Low** |
|  | Neither | No cases | No cases | No clinical cases | No neighbors | None detected | None detected |  |
| **Honduras** | **0** | **0** | **0** | **0** | **0** | **0** | **0** | **0- Low** |
|  | Neither | No cases | No cases | No clinical cases | No neighbors | None detected | None detected |  |
| **Nicaragua** | **0** | **0** | **0** | **0** | **0** | **0** | **0** | **0- Low** |
|  | Neither | No cases | No cases | No clinical cases | No neighbors | None detected | None detected |  |
| **El Salvador** | **0** | **0** | **0** | **0** | **0** | **0** | **0** | **0- Low** |
|  | Neither | No cases | No cases | No clinical cases | No neighbors | None detected | None detected |  |

HI: hemagglutination inhibition; NT: neutralization test; RT-PCR: Reverse transcription polymerase chain reaction; CHE: Current health expenditure

**Supplementary Table 3.**
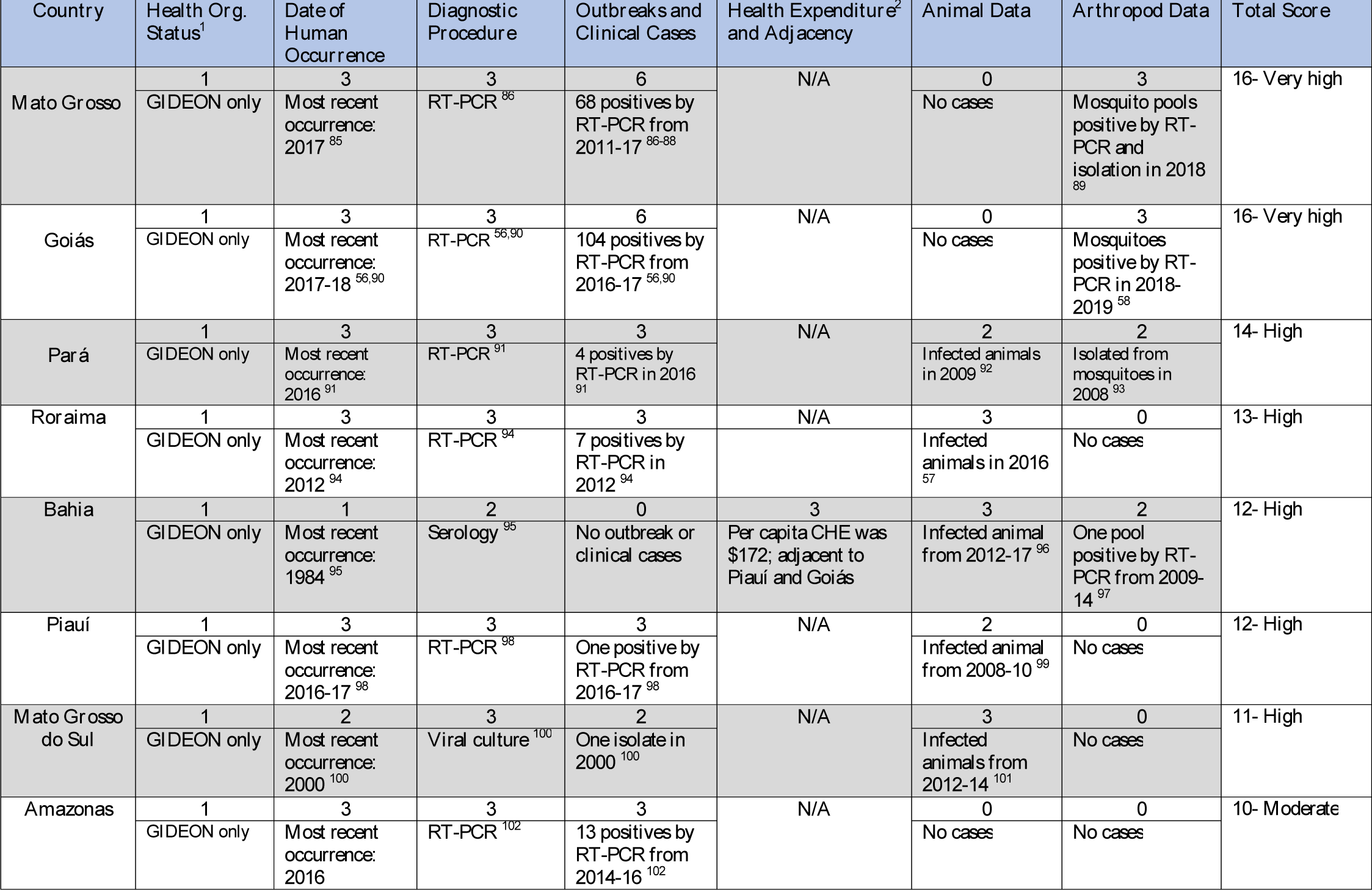
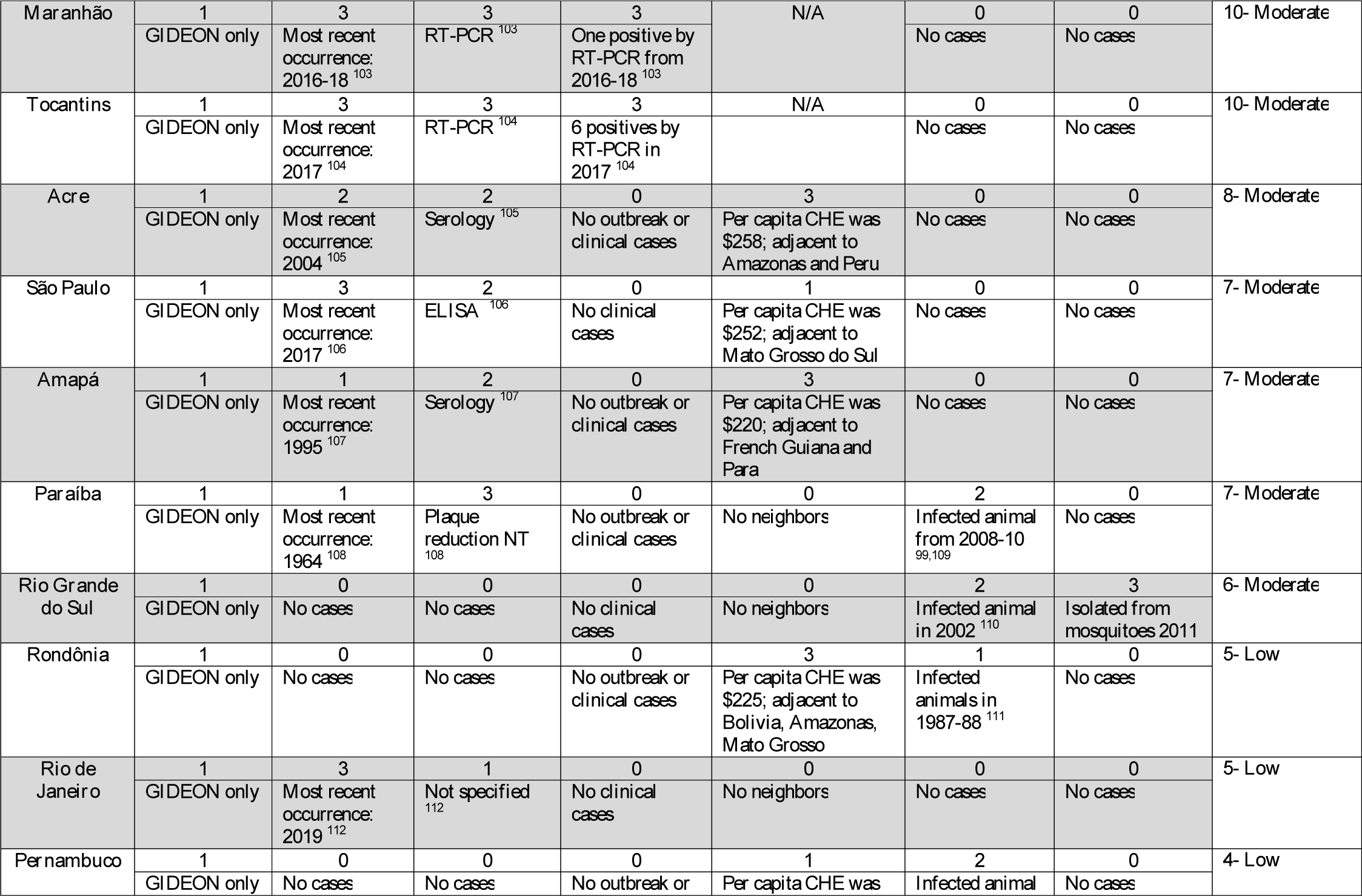
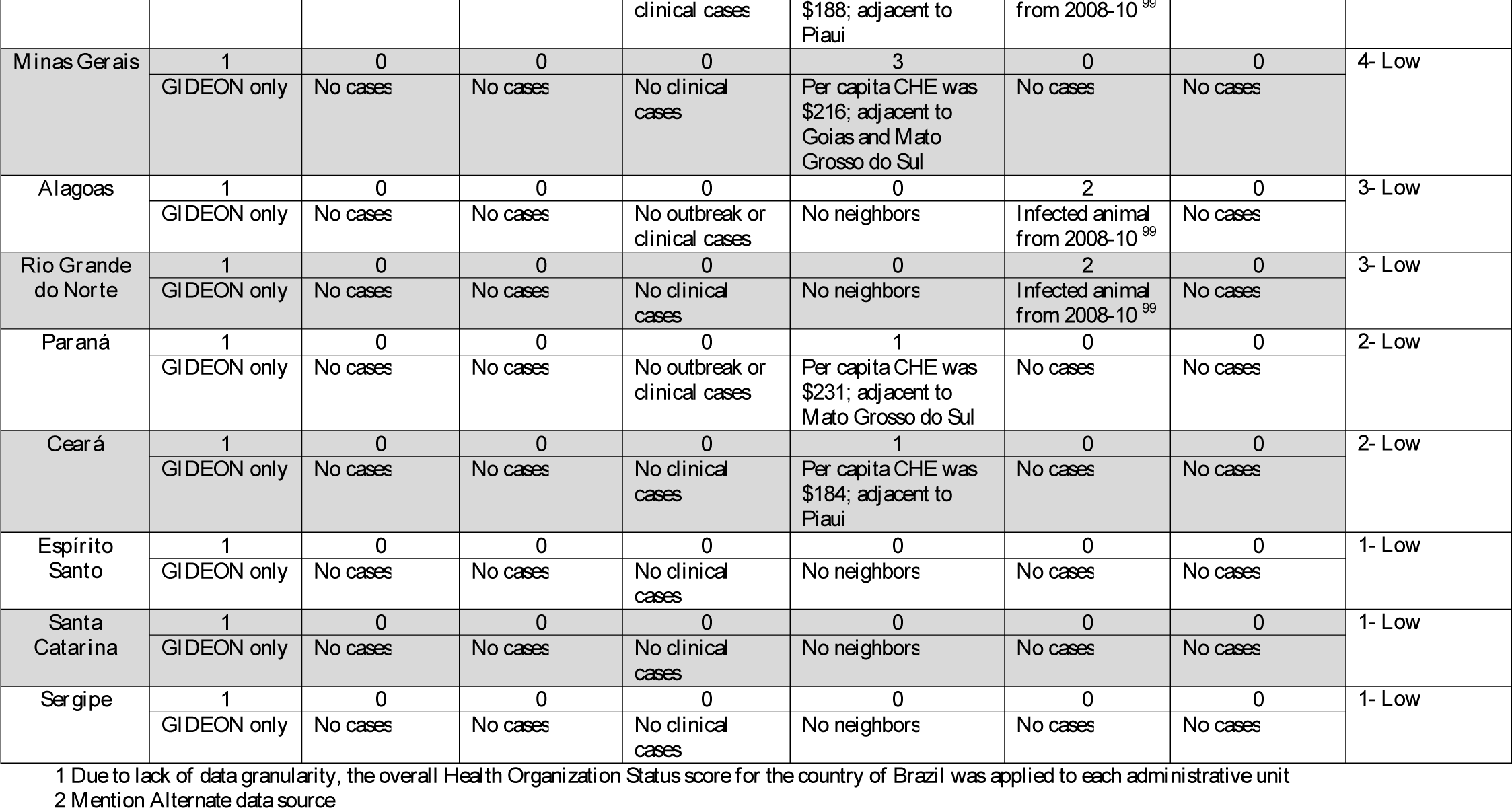
Evidence consensus by state (Brazil)

